# The impact of rapid near-patient STI testing on service delivery outcomes: a controlled interrupted time series study

**DOI:** 10.1101/2022.05.09.22274744

**Authors:** Scott R Walter, Joni Jackson, Gareth Myring, Maria Theresa Redaniel, Ruta Margelyte, Rebecca Gardiner, Michael D Clarke, Megan Crofts, Hugh McLeod, William Hollingworth, David Phillips, Peter Muir, Jonathan Steer, Jonathan Turner, Patrick J Horner, Frank de Vocht

**Author notes:** **Corresponding author:** Scott R Walter, National Institute for Health Research, Applied Research Collaboration West (NIHR ARC West), University Hospitals Bristol and Weston NHS Foundation Trust, 9th Floor, Whitefriars, Lewins Mead, Bristol, BS1 2NT, UK. joint last authors (Patrick J Horner, Frank de Vocht).

## Abstract

**Objectives:** To evaluate the impact of a new clinic-based rapid STI testing, diagnosis and treatment service on healthcare delivery and resource needs in a sexual health service.

**Design:** Controlled interrupted time series study.

**Setting:** Two sexual health services in UK: Unity Sexual Health in Bristol, UK (main site) and Croydon Sexual Health in London (control site).

**Participants:** Electronic patient records for all attendances during the period one year before and one year after the intervention.

**Intervention:** Introduction of an in-clinic rapid testing system for gonorrhoea and chlamydia in combination with revised treatment pathways.

**Outcome measures:** Time-to-test notification, staff capacity, cost per episode of care and overall service costs. We also assessed rates of gonorrhoea culture swabs, follow-up attendances, and examinations.

**Results:** Time-to-notification and the rate of gonorrhoea swabs significantly decreased following implementation of the new system. There was no evidence of change in follow-up visits or examination rates for patients seen in clinic related to the new system. Staff capacity in clinics appeared to be maintained across the study period. Overall, the number of episodes per week was unchanged in the Unity SHS, and the mean cost per episode decreased by 7.5% (95%CI 5.7%, 9.3%). Conclusions: The clear improvement in time-to-notification, while maintaining activity at a lower overall cost, suggests that the implementation of clinic-based testing in parallel to postal testing kits had the intended impact, which bolsters the case for more widespread rollout in SHS.

**Strengths and limitations of this study:**

- We used controlled interrupted time series models with confounder adjustment to estimate the effect of the intervention distinct from any background changes and independent of other time varying factors.
- Model validity was bolstered by using a relatively long time series with good temporal resolution.
- Data from both the main and control sites was derived from the same electronic patient record system.
- There was a general consensus between main and sensitivity analyses.
- Our study was limited by being non-randomised, having only one control site, and the follow up period for females being truncated by the impact of the Covid-19 pandemic.

## 1. Introduction

Sexually transmitted infection (STI) diagnoses are increasing in England with more than a 10% increase in new infections between 2016 and 2019[1]. Over the same period, a 19.2% increase in total consultations at sexual health services (SHS) was reported in England[2]. Open-access SHS providing rapid treatment and partner notification can reduce the risk of STI complications and infection spread[3,4,5]. Public Health England (now UK Health Security Agency) recommends that local SHS need to be available to both the general population and groups with greater sexual health needs[3]. Nevertheless, the central government’s public health grant, including SHS funding, has steadily decreased since 2015[6,7]. Despite diminishing resources, continued provision of SHS has been achieved through increased efficiencies at clinic-based services and introduction of online services[8,9].

Another approach to improving efficiency while ensuring quality, could be the introduction of near-patient testing (NPT) for chlamydia and gonorrhoea. Potential benefits include earlier diagnosis and treatment, reduced risk of sequelae and onward transmission, and reduction in unnecessary treatments, as well as reduced costs and clinician time due to reduction in gonorrhoea cultures, examinations and follow-up visits[10,11,12]. Although modelling studies suggest NPT can be cost-effective, this remains to be demonstrated in practice[10-14]. Research also suggests that reduced waiting times for STI test results may enhance patient acceptability[15,16] and increase testing uptake[17,18]. Importantly, patients have expressed preferences for earlier provision of results[19] due to the stress of waiting[20].

In November 2018, Unity Sexual Health (hereafter Unity), a UK specialist SHS, implemented a rapid nucleic acid amplification (NAAT) STI testing, diagnosis and treatment service for chlamydia and gonorrhoea, using the Hologic ‘Panther’ diagnostic platform in a clinic-based satellite laboratory [21]. It can deliver results in 3.5 hours by eliminating sample batching and transit times associated with microbiology laboratory testing.

We used a quantitative approach to evaluate the impact of the new rapid testing process on service delivery and resource needs of the Unity SHS.

## 2. Methods

### 2.1 Setting and design

This study is a quasi-experimental, controlled interrupted time series (CITS) design that used routinely collected electronic patient record (EPR) data. The intervention time points were defined differently for males and females: rapid STI testing was introduced on 12 November 2018 for males and 29 May 2019 for females. This study was approved by the Health Research Authority (South West) Research Ethics Committee, reference 18/SW/0090. Three members of the public who had used Unity services were involved in reviewing the proposed outcome measures and this informed study design.

### 2.2 Rapid STI service model

Eligibility criteria and treatment pathways differed for males and females. A graphical overview of each pathway is provided in the supplement (Figures S1 and S2) with pre-intervention pathway included for reference. Additional changes were made to the SHS related to staff capacity. Rapid STI asymptomatic consultations were reduced to 15 minutes, while the number of allocated patients per staff member for the walk-in clinic remained the same.

#### 2.2.1 Males

Male patients were eligible for the rapid STI pathway if they were asymptomatic or had urethritis symptoms. If asymptomatic, a brief history was taken prior to patient self-sampling for chlamydia and gonorrhoea and taking blood tests for HIV and syphilis. Men who have sex with men (MSM) were referred to a health adviser. Symptomatic men were asked to return four hours later when NAAT results were available. If positive, they received infection specific treatment; if negative a urethral smear was undertaken to diagnose non-gonococcal urethritis. Contacts of patients with gonorrhoea or chlamydia outside a two-week window were treated if NAAT-positive. Swabs for gonococcal culture and sensitivities were only taken after a NAAT-positive result for gonorrhoea or if gonococcal treatment was administered prior to the NAAT result.

#### 2.2.2 Females

Female asymptomatic patients without contraception needs were eligible for the rapid drop-off service. Women with abnormal vaginal discharge, not requiring bimanual or speculum examination to exclude pathology, self-swabbed and were treated on the results of microscopy and clinical findings at the time of visit and informed that chlamydia and gonorrhoea NAAT test results would be available within 4 8 hours. Trichomonas vaginalis (TV) culture was replaced with a more sensitive TV NAAT[22], also available within 48 hours. For contraceptive needs, a clinical consultation was necessary to determine the need for examination. A gonococcal culture swab was only taken after a NAAT-positive result for gonorrhoea or if gonococcal treatment was administered prior to NAAT result.

### 2.3 Control site

Croydon Sexual Health, a similar SHS in South London, was used as the control site to account for background changes unrelated to the intervention. This site has similar patient throughput (about 32,000 annual attendances compared to about 40,000 for Unity) and uses the same EPR system.

### 2.4 Data

Fully anonymised individual patient data extracted from the Unity and Croydon EPR systems[23] comprised demographic information, sexual behaviour, mode of presentation and attendances to the clinic, diagnostic testing and treatment. Analyses were based on a census of attendance level records.

Time-to-notification was defined from the text message notification system[23]. This included text message type for identifying test results messages, time stamps and anonymised patient identifiers. Numbers of NAAT postal testing kits were extracted from Unity’s records, while Croydon did not implemented these until after the study period.

Prior to analysis, data were checked for duplicates, implausible values and missingness. Individual variables were combined to generate indicator variables for complex cases, MSM, examinations, ethnic minority status. All time-related variables were derived from the date and time of each attendance.

For analysis, data were aggregated at weekly level over a two-year period centred at the intervention. For females, data were excluded from the first UK Covid-19-related lockdown (23 March 2020) due to changes in outcomes that could not be adequately accounted for in models. The study period for males was from 13 November 2017 to 10 November 2019, and for females 28 May 2018 to 22 March 2020.

### 2.5 Statistical analysis

There main study outcomes are detailed in Table 1. CITS models within a generalised linear modelling framework were applied to each outcome separately for males and females: ten models in total. Time was modelled as linear using consecutively numbered weeks, with time = 0 at the intervention point. A binary variable (period) representing pre- and post-intervention periods was defined by the respective male and female intervention dates.

**Table 1.** Definitions of main study outcomes.

| **Outcome measure** | **Definition** |
| --- | --- |
| 1. Rate of gonorrhoea culture swabs per consultation | Numerator: the number of GC swabs, urethral for male and cervical for female<br>Denominator: the number of consultations where these were defined as attendances for new, rebooked or walk-in patients |
| 2. Time to notification | Median time from sample collection until the patient was notified of the test result via text message |
| 3. Rate of examinations per symptomatic attendance | Numerator: the number of examinations of any type. This was based on a combination of variables used to record information about examinations (supplementary Table S1)<br>Denominator: all attendances where the patient was recorded as being symptomatic |
| 4. Rate of follow up attendances per episode of care | Numerator: the number of follow up attendances occurring within 30 days of an initial consultation<br>Denominator: the number of episodes involving at least 1 consultation |
| 5. Staff capacity – rate of patients seen per four-hour clinic | Numerator: number of patient consultations (any new, rebooked, walk-in or follow up attendance)<br>Denominator: number staff available for four-hour clinics |

Gonorrhoea culture swabs per consultation, follow-up attendances per care episode, examinations per symptomatic attendance and staff capacity were modelled as rates assuming a negative binomial distribution. These models generate rate ratios, presented as percentage changes. For time-to-notification, a normal distribution was assumed and results presented as differences in median time (days). This represents absolute measure of time including weekends as opposed to working days only.

The main variables in the models were *time, period* and site (Unity vs. Croydon) along with all two-way and three-way interactions, as per a CITS approach for estimating both a step change and slope change[24, 25]. Two key terms in the models represent intervention-related changes over and above any control site changes. The interaction *period x site* captures a differential step change for the intervention site compared to control site. While the three-way interaction term *time x period x site* captures different degrees of pre-post trend change for the intervention site compared to control site (supplement Figure S3).

Additional covariates were included in the models: proportions of complex patients, symptomatic patients and patients from an ethnic minority, plus mean patient age and calendar month. Since models of examination rate only analysed symptomatic patients, the proportion of symptomatic patients was excluded as a covariate. The proportion of MSM was only included in models for males. Complex cases were defined differently for males and females (definition S1).

Data for staff capacity was only available for Unity and was modelled as an uncontrolled interrupted time series spanning the duration of available denominator data: 1 January 2018 to 22 December 2019. The denominator could not be separated by gender, so this outcome was analysed for females and males combined, allowing two change points as per the respective intervention dates.

Where outcomes showed marked change over time, sensitivity analyses were conducted by fitting generalised additive models to account for potential non-linearity of trends. All analyses were conducted with the SAS System for Windows, version 9.4 (SAS Institute Inc.). Models were fitted using the GENMOD and GAM procedures.

### 2.6 Economic analysis

Postal testing kit data were combined with EPR data to estimate the total number of episodes per week (including those with negative postal tests and no clinic attendance). For estimating the difference in the mean number of episodes per week i) negative postal test episodes were assigned to weeks pro rata with asymptomatic episodes that included clinic attendance, and ii) the combined post-intervention analysis used data for the first 43 weeks only. Episode costs were estimated using unit costs of diagnostic tests provided by Unity SHS, and postal kit tests and staff time from the literature[12] inflated to 2021 values using a UK government GDP deflator[26]. Treatment costs were from the British National Formulary[27] (supplement Table S2). The cost of unreturned postal kits was allocated to episodes including a postal test result. Confidence intervals for differences in the number of episodes and cost per episode were calculated using the Normal approximation method.

## 3. Results

In the EHR Unity data, 48,776 attendances for females and 34, 413 for males were recorded during the study period, representing 32,482 and 22,073 episodes of care involving a clinic attendance, and 29,573 and 19,083 patients, respectively (Table 2). Patients were symptomatic in just over 20% of female attendances, and over 4 0% of male attendances. About 90% of female and 55% of male attendances were complex. Just over 30% of male attendances were by MSM.

**Table 2.** Summary of population characteristics and outcomes by site, gender and time period based on EPR data.

|  | Unity |  | Croydon |  |
| --- | --- | --- | --- | --- |
|  | Pre | Post | Pre | Post |
| **MALES** |  |  |  |  |
| Total attendances, n | 17626 | 16787 | 11920 | 12085 |
| Total episodes of care, n | 11445 | 10628 | 7946 | 8021 |
| Total patients, n | 9932 | 9151 | 6271 | 6335 |
| Symptomatic attendances, n (%) | 7307 (41.5%) | 7084 (42.2%) | 4735 (39.7%) | 4556 (37.7%) |
| Complex attendances, n (%) | 9869 (56.0%) | 9259 (55.2%) | 4458 (37.4%) | 4940 (40.9%) |
| Ethnic minority attendances, n (%) | 2834 (16.1%) | 3025 (18.0%) | 7244 (60.8%) | 7311 (60.5%) |
| MSM attendances, n(%) | 5300 (30.1%) | 5418 (32.3%) | 2529 (21.2%) | 2849 (23.6%) |
| Mean age, years | 30.2 | 30.8 | 34.9 | 35.1 |
| Urethral GC swabs per consultation | 0.18 | 0.11 | 0.08 | 0.07 |
| Median time to notification | 10.90 | 6.73 | 4.51 | 4.95 |
| Examinations per symptomatic attendance | 0.76 | 0.67 | 0.64 | 0.60 |
| Follow up attendances per episode | 0.40 | 0.36 | 0.50 | 0.37 |
| **FEMALES** |  |  |  |  |
| Total attendances | 28487 | 20289 | 20931 | 16910 |
| Total episodes of care | 18616 | 13866 | 13971 | 11660 |
| Total patients | 16779 | 12794 | 11799 | 9902 |
| Symptomatic attendances | 6312 (22.2%) | 4929 (24.3%) | 6860 (32.8%) | 5561 (32.9%) |
| Complex attendances | 26022 (91.3%) | 18173 (89.6%) | 12328 (58.9%) | 11221 (66.4%) |
| Ethnic minority attendances | 3979 (14.0%) | 3067 (15.1%) | 12647 (60.4%) | 10107 (59.8%) |
| Mean age | 25.1 | 25.8 | 29.8 | 30.4 |
| Cervical GC swabs per consultation | 0.20 | 0.04 | 0.03 | 0.03 |
| Median time to notification (median, IQR) | 10.58 | 3.52 | 4.90 | 5.32 |
| Examinations per symptomatic attendance | 0.73 | 0.70 | 0.58 | 0.60 |
| Follow up attendances per episode | 0.36 | 0.34 | 0.31 | 0.23 |

### 3.1 Males

There were significant changes in the rate of gonorrhoea culture swabs for males associated with the intervention. A small increase at the time of the intervention for Unity (+6.5%) compared to a large decrease for Croydon (−43.7%), resulted in a significant adjusted step-increase for Unity (+89.1%, 95% confidence interval [CI] +37.1%, +160.6%, p<0.001) (Table 3 and Figure 1A). However, this was not observed in the sensitivity analysis allowing for non-linear trends (supplement Table S3 and Figure S4 A). This was followed by a significant adjusted downward change in post-intervention trend of -3.2% per week (95% CI -4 .3%, -2.1%, p<0.001). The long-term result of these two effects was an overall decrease from 35-50 swabs per week, pre-intervention, to below 10 at the end of the study period, translating to 849 swabs avoided over the post-intervention period.

**Table 3.** Intervention-related model estimates for females and males.

| Outcome | Change at time of intervention |  |  | Trend change following intervention |  |  |
| --- | --- | --- | --- | --- | --- | --- |
|  | *Intervention series* | *Control series* | *Intervention vs. control, % change (95% CI)* | *Intervention series* | *Control series* | *Intervention vs. control, % change per week (95% CI)* |
| **MALES – 12<sup>th</sup> November 2018** |  |  |  |  |  |  |
| **1. Gonorrhoea culture swabs per consultation** | +6.5% | -43.7% | **+89.1% (+37.1%, +160.9%)** | -3.6% | -0.3% | **-3.2% (-4.3%, -2.1%)** |
| **2. Time to notification** | +2.2 days | +5.8 days | **+3.6 (+1.7, +5.5) days** | -0.19 days | +0.03 days | **-0.2 (-0.3, -0.2) days** |
| **3. Examinations per symptomatic attendance** | +3.6% | -1.6% | +5.4% (-7.5%, +20.0%) | -0.21% | -0.16% | -0.04% (-0.5%, +0.4%) |
| **4. Follow up attendances per episode** | -9.0% | -11.9% | +3.3% (-14.6%, +24.9%) | +0.23% | -0.001% | +0.30% (+0.31%, +0.96%) |
| **FEMALES – 29<sup>th</sup> May 2019** |  |  |  |  |  |  |
| **1. Gonorrhoea culture swabs per consultation** | -38.7% | +3.6% | **-40.8% (-61.6%, -8.8%)** | -6.1% | -0.1% | **-6.1% (-7.8%, -4.5%)** |
| **2. Time to notification** | -2.5 days | -0.4 days | -2.1 (-4.5, 0.3) days | -0.11 days | -0.0001 days | **-0.1 (-0.2, -0.0) days** |
| **3. Examinations per symptomatic attendance** | -1.3% | -2.2% | +1.0% (-11.4%, +15.1%) | +0.09% | +0.03% | +0.1% (-0.4%, +0.5%) |
| **4. Follow up attendances per episode** | -8.2% | +2.7% | -10.6% (-27.6%, +10.3%) | -0.42% | +0.22% | -0.64% (-1.41%, +0.14%) |

Note: Results for outcome 5 (staff capacity) reported separately in the text.

**Figure 1.**
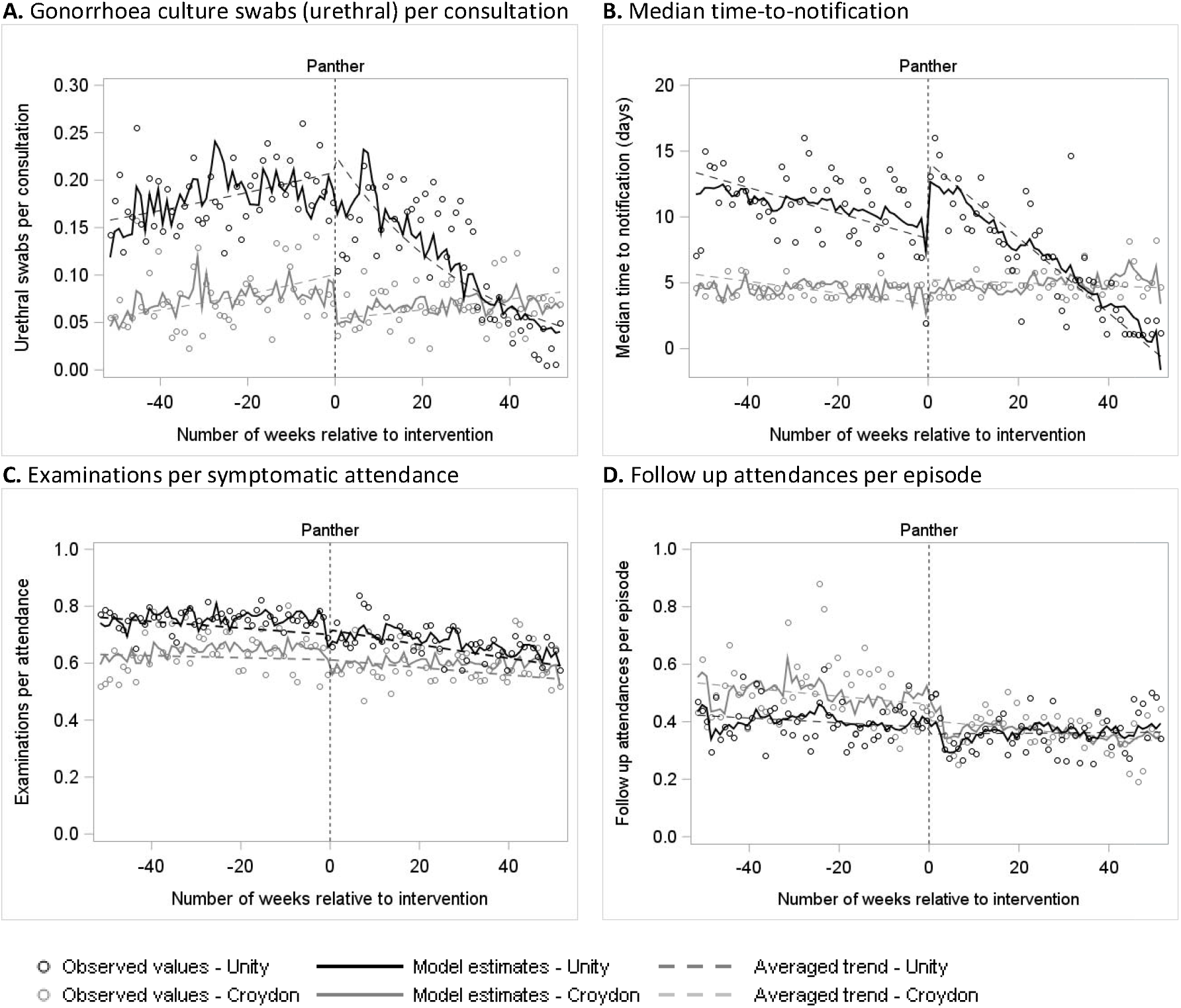
Modelled outcome estimates for males. ‘Panther’ indicates the intervention-date representing the first week the Panther system was implemented for the male pathway: 12 November 2018.

Time-to-notification increased by an estimated 3.6 days (95% CI 1.7, 5.5 days, p<0.001) at the time of the intervention, relative to controls, and a similar increase was observed in the sensitivity analysis. However, this was followed by an overall long-term decrease of -0.2 days per week (95% CI -0.3, -0.2 days, p<0.001) through the post-intervention period. That is, the pre-intervention weekly median of around eight to nine days dropped to around 2 days after the Panther system had been in place for a year (Figure 1B).

We found no evidence of a meaningful change in rates of examinations or follow-up attendances associated with the intervention.

### 3.2 Females

For females, there was significant decrease in the rate of gonorrhoea culture (GC) swabs: -40.8% (95% CI -61.6%, -8.8%, p=0.02) at the time of intervention, adjusted for control changes (Table 3, Figure 2A). This was followed by a significant decrease in trend through the post-intervention period, with an adjusted change of -6.1% per week (95% CI -7.8%, -4.5%, p<0.001). These changes represent a decrease from an estimated 0.22 swabs per consultation (over 30 swabs per week) immediately before the intervention to 0.14 immediately after (20 to 25 per week) and down to 0.01 at the end of the study period (less than five per week). Over the 43-week post-intervention period, an estimated 1542 swabs were avoided.

**Figure 2.**
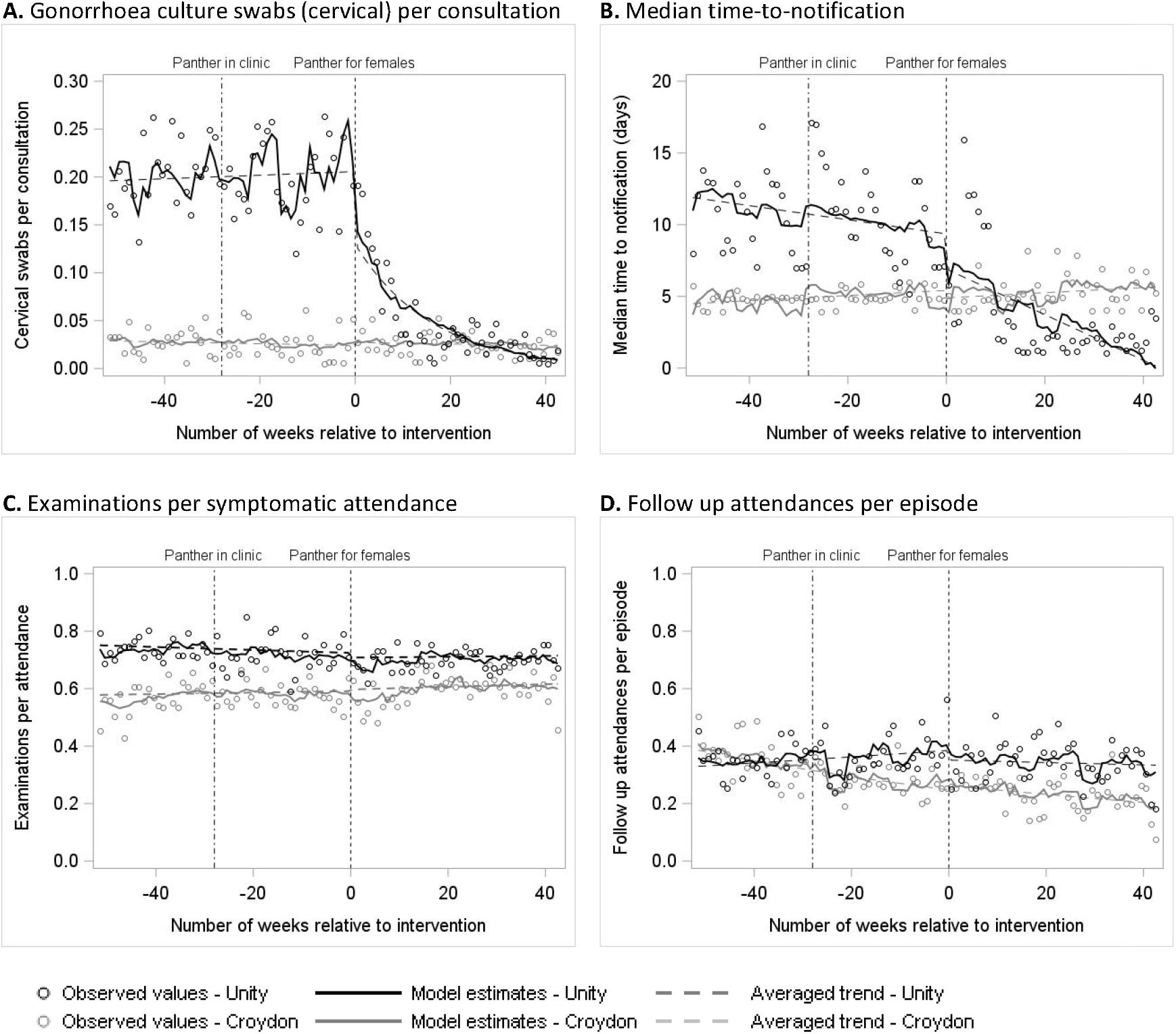
Modelled outcome estimates for females. ‘Panther for females’ indicates the intervention-date representing the first week the Panther system was implemented for the female pathway: 29 May 2019.

For time-to-notification, there was some evidence of a decrease of 2.1 days (95% CI -4.5, 0.3 days, p=0.08) at the time of the intervention, adjusted for the control group, although this estimate does not rule out chance. There was stronger evidence of a downward change in trend, estimated at -0.1 days per week (95% CI -0.20, -0.0 days, p=0.01) over the post-intervention period. To illustrate, the estimated median time-to-notification was eight to nine days just before the intervention, but a year later had dropped to around one day.

### 3.3 Staff capacity

The main analysis of staff capacity showed a significant trend change at the time of the male intervention (−1.1% per week, 95%CI -1.7%, -0.5%, p<0.001) and a significant step change at the time of the female intervention (+14.3%, 95% CI +3.4%, +26.3%, p=0.009) (Figure 3). However, the sensitivity analysis showed step changes in the opposite direction to the main analysis (supplement Figure S6), suggesting inconclusive evidence of change.

**Figure 3.**
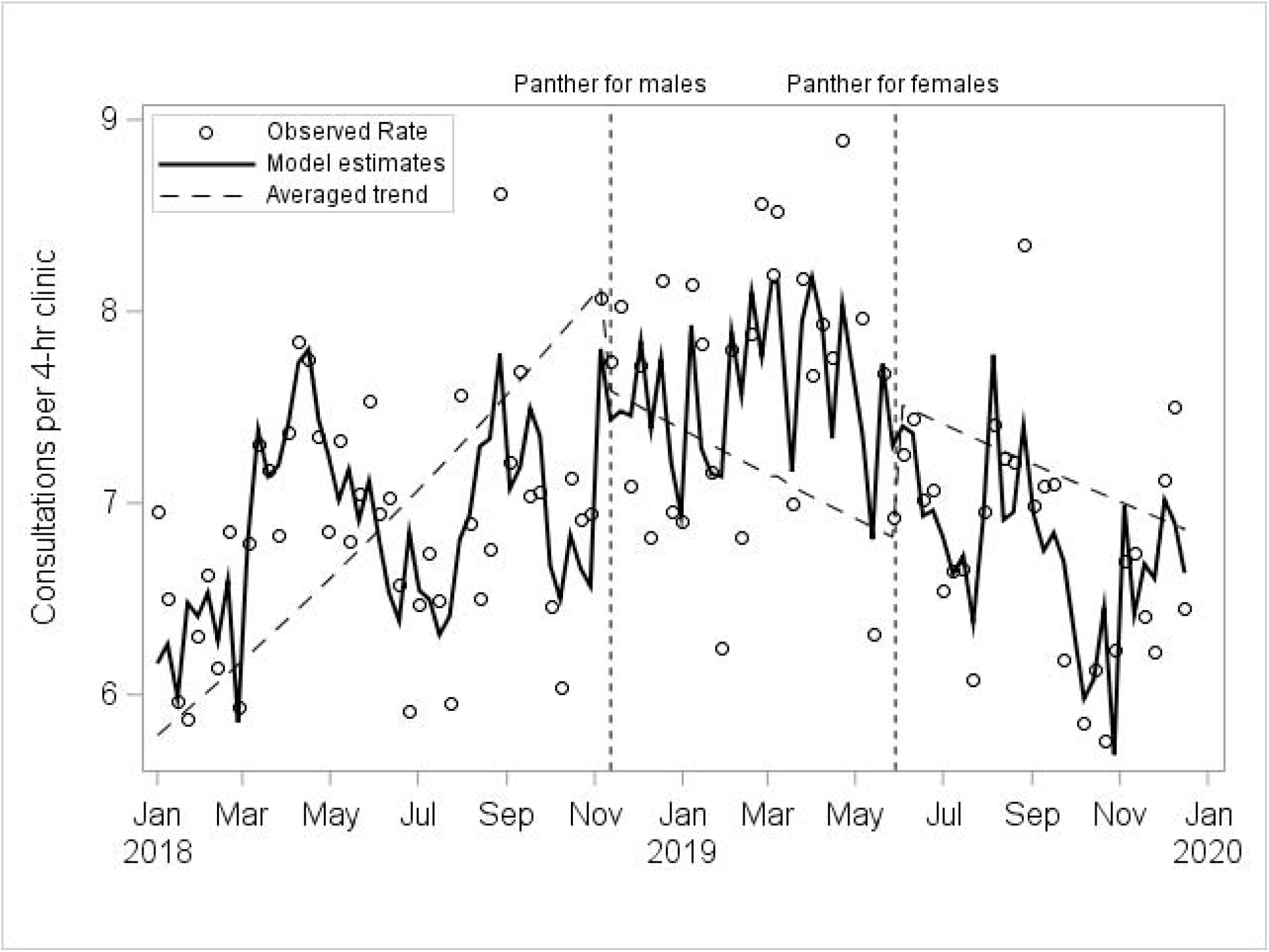
Modelled estimates of staff capacity for males and females combined.

### 3.4 Episodes and costs

Overall, the Unity SHS experienced a substantial increase in the weekly number of asymptomatic negative episodes managed via postal test kits, particularly for males, while both asymptomatic negative episodes seen in the clinic and symptomatic episodes decreased (Table 4). The mean cost per symptomatic episode increased by 9.2% to £69.04, while this was outweighed by a decrease of 13.5% to £26.23 for costs per asymptomatic episode, resulting in a combined decrease of 7.5%. The total cost per week decreased by 4 .7%, largely due to the reduction in both the number and cost of episodes for asymptomatic females who attended the clinic.

**Table 4.** Unity clinic pre- and post-intervention estimates of mean number of episodes per week, mean cost per episode and mean cost per week.

|  | Male |  |  |  |  | Female |  |  |  |  | Total |  |  |  |  |
| --- | --- | --- | --- | --- | --- | --- | --- | --- | --- | --- | --- | --- | --- | --- | --- |
|  | pre* | post* | % change | 95% CI |  | pre* | post** | % change | 95% CI |  | pre* | post** | % change | 95% CI |  |
| *Mean number per week* |  |  |  |  |  |  |  |  |  |  |  |  |  |  |  |
| Asymptomatic | 190.2 | 223.1 | 17.3 | 9.5 | 25.1 | 356.2 | 350.7 | -1.5 | -7.9 | 4.9 | 546.3 | 573.4 | 5.0 | 0.0 | 9.9 |
| Postal negatives | 70.5 | 111.5 | 58.2 | 48.7 | 67.7 | 96.3 | 124.9 | 29.7 | 22.3 | 37.0 | 166.8 | 236.2 | 41.6 | 35.7 | 47.4 |
| Other^ | 119.6 | 111.5 | -6.8 | -13.7 | 0.2 | 259.9 | 225.9 | -13.1 | -19.2 | -7.0 | 379.5 | 337.2 | -11.1 | -15.8 | -6.5 |
| Symptomatic | 92.7 | 85.0 | -8.3 | -13.9 | -2.7 | 84.4 | 77.8 | -7.8 | -14.6 | -1.1 | 176.7 | 163.4 | -7.5 | -11.8 | -3.2 |
| Total | 282.8 | 308.0 | 8.9 | 2.6 | 15.2 | 440.2 | 429.0 | -2.5 | -8.7 | 3.6 | 723.0 | 736.8 | 1.9 | -2.5 | 6.3 |
| *Cost per episode (£)* |  |  |  |  |  |  |  |  |  |  |  |  |  |  |  |
| Asymptomatic | 36.47 | 30.92 | -15.2 | -19.1 | -11.3 | 27.04 | 24.23 | -10.4 | -13.3 | -7.5 | 30.31 | 26.23 | -13.5 | -15.9 | -11.0 |
| Symptomatic | 63.09 | 69.56 | 10.3 | 6.7 | 13.8 | 63.36 | 67.65 | 6.8 | 4.3 | 9.2 | 63.22 | 69.04 | 9.2 | 6.9 | 11.5 |
| Total | 45.19 | 41.58 | -8.0 | -10.8 | -5.2 | 33.98 | 32.14 | -5.4 | -7.7 | -3.1 | 38.36 | 35.47 | -7.5 | -9.3 | -5.7 |
| *Cost per week (£)* |  |  |  |  |  |  |  |  |  |  |  |  |  |  |  |
| Resource |  |  |  |  |  |  |  |  |  |  |  |  |  |  |  |
| Postal kit | 382 | 592 | 55.0 | 45.9 | 64.1 | 629 | 848 | 34.8 | 27.4 | 42.2 | 1010 | 1437 | 42.3 | 36.5 | 48.1 |
| In clinic diagnostic test | 1962 | 1886 | -3.9 | -9.8 | 2.1 | 1452 | 1213 | -16.5 | -22.9 | -10.1 | 3413 | 3155 | -7.6 | -11.9 | -3.3 |
| Consultation staff time | 7497 | 7349 | -2.0 | -7.3 | 3.4 | 9396 | 8583 | -8.7 | -15.0 | -2.3 | 16893 | 15959 | -5.5 | -9.5 | -1.5 |
| Treatment | 3024 | 2896 | -4.2 | -13.1 | 4.6 | 3534 | 3085 | -12.7 | -20.3 | -5.1 | 6558 | 6014 | -8.3 | -14.4 | -2.2 |
| Symptom status |  |  |  |  |  |  |  |  |  |  |  |  |  |  |  |
| Asymptomatic | 6949 | 6883 | -1.0 | -8.5 | 6.6 | 9673 | 8448 | -12.7 | -18.9 | -6.4 | 16622 | 15392 | -7.4 | -12.3 | -2.5 |
| Symptomatic | 5915 | 5840 | -1.3 | -7.5 | 5.0 | 5338 | 5280 | -1.1 | -8.7 | 6.6 | 11253 | 11174 | -0.7 | -5.5 | 4.0 |
| Total | 12865 | 12723 | -1.1 | -6.7 | 4.5 | 15010 | 13728 | -8.5 | -14.4 | -2.6 | 27875 | 26565 | -4.7 | -8.6 | -0.8 |

\* based on 52 week period.
\*\* based on 43 week period
^ includes positive postal test kits

## 4. Discussion

We have quantitatively evaluated the impact of a first-of-its-kind integrated rapid STI testing on service delivery. Previous NPT assessments have taken a mathematical modelling approach[11-13]. The only other direct assessment of a chlamydia and gonorrhoea NPT in practice related to a rapid testing service model for asymptomatic patients[28]. This is the first study to quantify the effect of rapid chlamydia and gonorrhoea NPT on gonorrhoea culture swabs, time-to-notification, examinations, follow-up visits, staff capacity, and costs.

The substantial long term post-intervention decrease in the rate at which gonorrhoea swabs were sent for culture, for both males and females, was expected to some extent since patients with negative rapid tests in the new pathway avoided the need for cultures. Adams et al. [11] identified reduced gonorrhoea cultures as a key part of NPT-related cost reduction, although there has been no direct or simulated assessment of expected change in the number of cultures.

The trajectory of the decline in gonorrhoea swab rates following the intervention differed between males and females. The sensitivity analysis capturing non-linear trends suggested substantial decreases for males began more than six months after the intervention, with the lowest rates at one year post-intervention (Figure S4A). In contrast, rates for females appeared to respond to the intervention almost immediately and stabilise at a much lower level within about six months (Figure S5A). The differing implementation timeframes may reflect several barriers to implementation with the initial rollout for males, including providing training to a large group staff with varying timetables exacerbated by understaffing and budget cuts; variable application of eligibility criteria for the new service; and iterative revision of the new system and pathway[29]. There may also have been some just-in-case culture testing in the early stages until staff confidence in the system was established. With these issues largely resolved when the system was implemented for females, the transition appeared both smoother and faster, and this concurs with staff experience.

We estimated that median time-to-notification decreased from more than a week down to one or two days over the post-intervention period. However, given that it was not possible to separate out all rapid test results (e.g. notifications labelled “all negative”) and that we estimated real time rather than working days, the median time was likely lower, particularly for positive results. This is broadly consistent with findings from Whitlock et al. [28] who reported an average time-to-notification of 0.27 days for a new rapid NAAT testing service compared to 8.95 days for an off-site testing service for symptomatic patients.

The temporary increase in median time-to-notification for males after the intervention may result from the implementation challenges outlined above[29] in addition to a clinician-reported backlog in the early stages of transitioning to the new system. Once again, for males the transition appeared to take place over the full post-intervention period, while the equivalent period for females appeared faster with the lowest post-intervention sensitivity estimates occurring 21 weeks after the new system was implemented (supplement Figures S4B and S5B).

We observed no clear evidence of intervention-related changes in rates of examinations, follow up visits or staff capacity. All three were necessarily constructed from combinations of variables as there was no dedicated data field for each in the data. Although we did not detect a positive change, it is important to note that there was no evidence of a deleterious impact of the rapid testing service on any of these outcomes.

Staff capacity showed some evidence of intervention-related change, although the rate of patients seen per four-hour clinic was at similar levels at the end of the study period as at the start. For asymptomatic patients, the provision of postal testing kits and the introduction of shorter appointments more than likely increased staff capacity for this subgroup. It also reduced the queueing time for walk-in clinics. Conversely, case-mix in the walk-in clinics became more demanding, with patients more likely to be symptomatic and/or complex[29], which may explain the lack of observed improvement in staff capacity during clinics. The lack of evidence for a capacity decrease through the implementation period despite a more demanding patient group and the growing numbers of asymptomatic patients being tested both suggest increased capacity of the SHS overall.

The change in management of asymptomatic clinical attendances, supported by the existing postal testing kit system, was a key component of the overall cost reduction following the introduction of the Panther technology, with decreases in both mean cost per asymptomatic episode (13.5%) and weekly asymptomatic costs (7.4%). Although the cost of symptomatic episodes increased, consistent with the reported increase in complexity of symptomatic patients in clinic, this was counteracted by a reduction in the number of weekly symptomatic attendances.

### 4.1 Strengths and limitations

We conducted a prospective real-time evaluation of a large integrated rapid STI service. We used a CITS framework with both a control site and confounder adjustment to estimate the effect of the intervention distinct from any background changes and independent of other time varying factors. This was bolstered by using a relatively long time series with good temporal resolution. The robustness of our analysis was supported by both sites using the same EPR system and the general consensus between main and sensitivity analyses.

In light of the target trial framework for natural experiments[30], our study was limited by being non-randomised, having only one control site, relying on the construction of certain outcomes from multiple variables, and the impact of the Covid-19 pandemic on the follow up period for females. The unit costs were based on data provided by Unity SHS and estimates from literature, and commissioners will need to assess their applicability to their locality.

### 4.2 Implications and conclusions

Several studies have suggested that NPT benefits include earlier diagnosis and treatment, reduced risk of sequelae and onward transmission, reduction in unnecessary treatments, earlier partner notification and reduced anxiety [10,28].

This quantitative assessment of the first UK implementation of rapid chlamydia and gonorrhoea testing within an integrated service revealed clear benefits, namely: reduced gonorrhoea culture swabs and shortened time-to-notification. These improvements, while maintaining activity at a lower overall cost, suggests that the introduction of clinic-based rapid testing had the intended impact, and this is in line with previous NPT modelling studies [10,11]. The qualitative evaluation of this rapid STI service also reported that patients valued faster results and avoiding unnecessary treatment, and that the better targeting of infection-specific treatment improved antimicrobial stewardship[29].

These results provide real-life evidence to support the benefits of a rapid testing service anticipated by modelling studies and strengthen the case for more widespread rollout in SHS.

## Supporting information

Definition S1

## Data Availability

Our data sharing agreement with the data controllers prohibits sharing data extracts outside of the University of Bristol research team.

## Acknowledgements

The authors would like to thank Ed Hulse at Mill Systems for his indispensable assistance with the data extracts.

## Author statement

PH, MTR, FdV and HM conceptualized the evaluation; MTR and FdV are quantitative evaluation leads; WH and HM are health economic evaluation leads; SW, JJ, RM and MTR acquired the analysis datasets; SRW conducted the effectiveness analysis with supoprt from JJ, RM, MTR, PH and FdV; GM conducted the cost-effectiveness analysis with support from HM and WH; RG, MDC, MC, DP, PM, JS and JT advised on the study methodology, analysis and interpretation of results; SRW wrote the initial draft of the manuscript; all authors reviewed and edited the manuscript for content and approved the submission.

### Data sharing statement

Anonymised individual-level data for this study comes from the electronic patient record system of the Unity Sexual Health and Croydon Sexual Health services (data controllers). Our data sharing agreement with the data controllers prohibits sharing data extracts outside of the University of Bristol research team. The data is available upon request from the data controllers. Copies of the study protocol are available on available on the University of Bristol’s institutional repository: http://hdl.handle.net/1983/79f8f4a6-6710-42e3-b1fd-4fc836ea7618

### Licence statement

I, the Submitting Author has the right to grant and does grant on behalf of all authors of the Work (as defined in the below author licence), an exclusive licence and/or a non-exclusive licence for contributions from authors who are: i) UK Crown employees; ii) where BMJ has agreed a CC-BY licence shall apply, and/or iii) in accordance with the terms applicable for US Federal Government officers or employees acting as part of their official duties; on a worldwide, perpetual, irrevocable, royalty-free basis to BMJ Publishing Group Ltd (“BMJ”) its licensees and where the relevant Journal is co-owned by BMJ to the co-owners of the Journal, to publish the Work in BMJ Open and any other BMJ products and to exploit all rights, as set out in our licence.

The Submitting Author accepts and understands that any supply made under these terms is made by BMJ to the Submitting Author unless you are acting as an employee on behalf of your employer or a postgraduate student of an affiliated institution which is paying any applicable article publishing charge (“APC”) for Open Access articles. Where the Submitting Author wishes to make the Work available on an Open Access basis (and intends to pay the relevant APC), the terms of reuse of such Open Access shall be governed by a Creative Commons licence – details of these licences and which Creative Commons licence will apply to this Work are set out in our licence referred to above.

