## Supplementary material for "The impact of rapid near-patient STI testing on service delivery outcomes: a controlled interrupted time series study": Figure S1

**Figure S1.** Male treatment pathway before (usual care pathway) and after (rapid pathway) implementation of the Panther rapid results system. Reproduced from: Lorenc A, Kesten J, Brangan E, Horner PJ, Clarke M, Crofts M, Turner J, Muir P, Horwood J. What can be learnt from a qualitative evaluation of implementing a rapid sexual health testing, diagnosis and treatment service? BMJ Open, 2021; 11: e050109. doi: 10.1136/bmjopen-2021-050109.

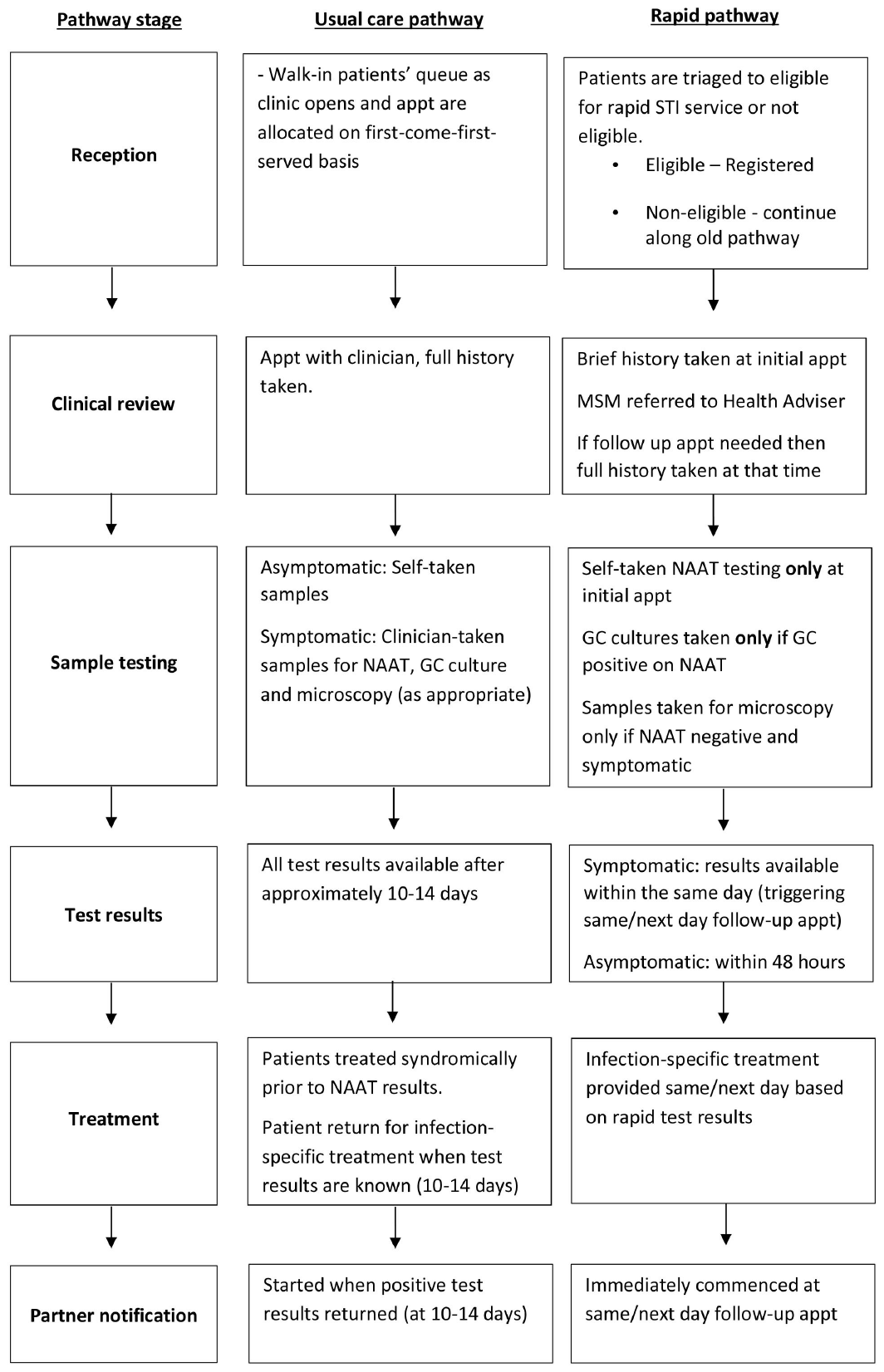
