## Supplementary material for "The impact of rapid near-patient STI testing on service delivery outcomes: a controlled interrupted time series study": Figure S2

**Figure S2.** Overview of female rapid treatment pathway for asymptomatic and symptomatic patients before (usual care pathway) and after (rapid pathway) implementation of the Panther rapid results system. NAAT = Nucleic Acid Amplification Test GC = Gonorrhoea CT= Chlamydia, TV= *Trichomonas vaginalis*

**Pathway stage**

Walk-in patients’ queue as clinic opens and appointments are allocated on first-come-first-served basis.

**Reception**

Patients triaged to rapid STI service or not:

● Symptomatic and Eligible = Registered

● Symptomatic and non-eligible = modified usual care pathway (GC/CT and TV NAAT results <48 hrs) GC cultures only taken if GC NAAT positive or patient receives treatment for GC before results of GC NAAT available

**Rapid pathway: asymptomatic**

**Usual care pathway**

**Rapid pathway: symptomatic**

Patients are triaged to rapid STI service or not:

● Asymptomatic and Eligible = Registered

● Asymptomatic and Non-eligible = continue along modified usual care pathway

**Treatment**

Immediately commenced at same/next day follow-up appt.

Immediately commenced at same/next day follow-up appt.

**Partner notification**

Started when positive test results returned (at 10-14 days).

Infection-specific treatment provided same/next day based on rapid test results.

Patients treated syndromically prior to NAAT results.

Patients return for infection-specific treatment when test results are known (10-14 days).

Infection-specific treatment provided same/next day based on rapid test results.

**Treatment**

**Test results**

All test results available after approximately 10-14 days.

Results available within 48 hours.

Results of vaginal microscopy available in 20 mins and patient is then reviewed by clinician CT/GCTV NAAT results available within 48 hours

Self-taken:

● Vaginal CT/GC/TV NAAT

● Vaginal swab for Gram staining

GC cultures taken ***only*** if GC positive on NAAT or if treatment as GC contact within 2 weeks of sexual intercourse with contact.

Self-taken NAAT testing ***only*** at initial appt

GC cultures taken ***only*** if GC positive on NAAT or if treatment as GC contact within 2 weeks of sexual intercourse with contact

**Sample testing**

● Asymptomatic: Self-taken samples

● Symptomatic: Clinician-taken samples for CT/GC NAAT. If vaginal symptoms or pelvic pain then speculum exam and swabs taken for microscopy, TV and GC culture (as appropriate).

History taken and no indications for visual inspection or pelvic examination required.

If follow up appt. needed then full history taken at that time

**Clinical review**

Appt. with clinician, full history taken.

Brief history taken at initial appt.

If follow up appt. needed then full history taken at that time
