## Supplementary material for "The impact of rapid near-patient STI testing on service delivery outcomes: a controlled interrupted time series study": Table S1

**Table S1.** Definition of examination of any type based on a combination of two examination-related variables.

| **Categorical exam variable** | **Free text exam variable** | **Define as exam – MALES** | **Define as exam - FEMALES** |
| --- | --- | --- | --- |
| Yes | Notes indicating exam | Yes | Yes |
| Yes | Missing | Yes | Yes |
| No | Notes indicating exam | Yes | Yes |
| No | Notes indication NO exam |  |  |
| No | Missing |  |  |
| External only | Notes indicating exam | Yes | Yes |
| External only | Notes indication NO exam |  |  |
| External only | Missing |  | Yes |
| Speculum and external | Notes indicating exam | Yes | Yes |
| Speculum and external | Notes indication NO exam |  |  |
| Speculum and external | Missing |  | Yes |
| Missing | Notes indicating exam | Yes | Yes |
| Missing | Notes indication NO exam |  |  |
| Missing | Missing |  |  |

Notes: The categorical exam variable was intended for use with female patients but was sometimes used for males.
