## Supplementary material for "The impact of rapid near-patient STI testing on service delivery outcomes: a controlled interrupted time series study": Figure S3

**Figure S3.** Diagrams to illustrate controlled interrupted time series variables for estimating A) changes at the time of intervention and B) changes in trends.

**A. Changes at the time of intervention**

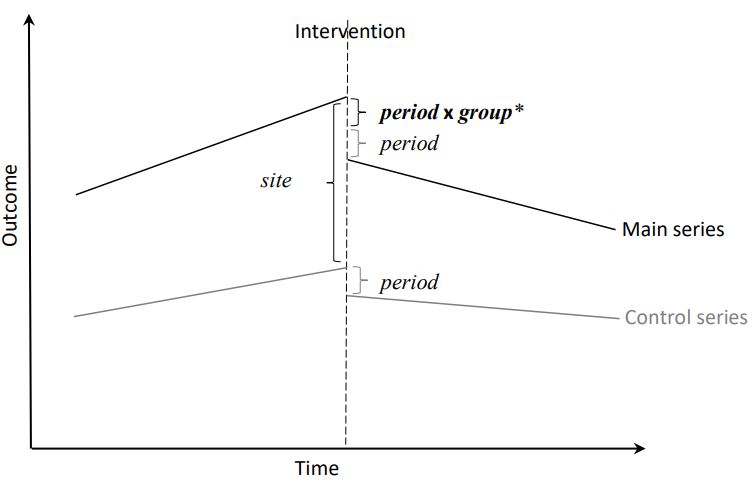

**B. Trend changes**

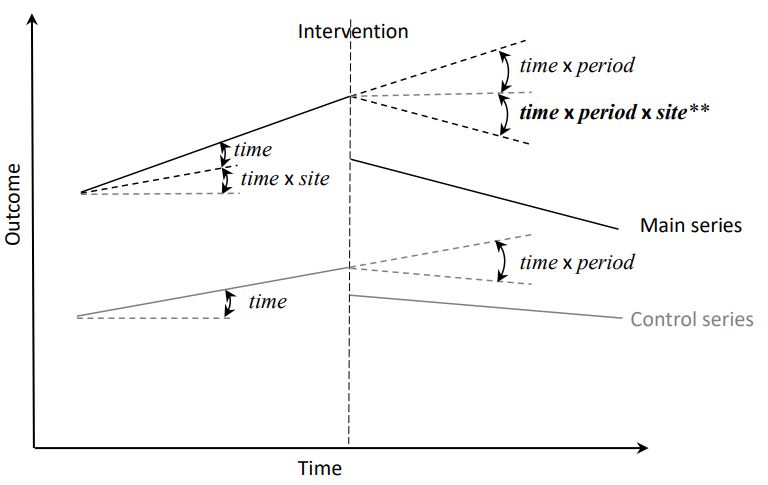

Note: *site* is a binary variable indicating either intervention or control sites; *period* is also binary indicating pre- or post-intervention periods; *time* is a continuous variable consecutively numbering each time unit (weeks in this study) with *time*=0 centred at the intervention

* *period* x *site* represents change in the intervention site at the time of intervention over and above any changes in the control site

** *time* x *period* x *site* represents change in trend for the intervention site over and above any trend changes in the control site
