## Supplementary material for "The impact of rapid near-patient STI testing on service delivery outcomes: a controlled interrupted time series study": Table S2

**Table S2.** Unit costs.

Unit costs of postal kit tests, and staff time[12] were from the literature and inflated to 2021 values using a UK government GDP deflator[26]. Local unit costs of diagnostic tests were provided by Unity SHS. Treatment costs were from the British National Formulary[27].

| **Resource use** | **Unit cost** |
| --- | --- |
| **Tests** |  |
| Lab CT/GC test | £8.10 |
| POCT CT/GC | £9.48 |
| GC culture swab | £6.13 |
| Male postal kit returned | £4.44 |
| Male postal kit not returned | £3.61 |
| Female postal kit returned | £4.08 |
| Female postal kit not returned | £3.24 |
| **Consultation staff time** |  |
| Follow-up | £9.33 |
| Male non-complex | £29.03 |
| Male complex | £46.54 |
| MSM complex | £42.97 |
| Female non-complex | £29.03 |
| Female complex | £52.26 |
| **Treatment** |  |
| 13.5mg Levonorgestrel IUS | £69.22 |
| 6mg norelgestromin and 600micrograms ethinylestradiol | £19.51 |
| Aciclovir 400 mg (tds for 5 Days) | £0.79 |
| Aciclovir 400mg (bd for 6 months) | £17.64 |
| Aciclovir 400mg (bd for 3 months) | £8.82 |
| Aciclovir 800mg (tds for 2 days) | £0.59 |
| Amoxicillin 250 mg tds for 5 days | £0.98 |
| Amoxicillin 500mg | £1.01 |
| Anusol Cream | £2.49 |
| Anusol Ointment | £2.49 |
| Anusol Suppositories | £1.74 |
| Aqueous Cream BP 100g Tube | £0.77 |
| Aqueous Cream BP 500g Tub | £3.85 |
| Azithromycin 1g (2 x 500mg tablets) | £0.81 |
| Azithromycin 1g (4 x 250mg capsules) | £1.24 |
| Azithromycin 1g stat, then 500mg od for 2 days | £1.21 |
| Azithromycin 1g stat, then 500mg od for 4 days | £2.42 |
| Azithromycin 2g o stat | £1.62 |
| Benzathine Benzylpenicillin 2.4 million units on day 0 | £9.50 |
| Benzathine benzylpenicillin 2.4 million units at day 7 | £9.50 |
| Benzathine benzylpenicillin 2.4 million units at day14 | £9.50 |
| Betamethasone Valerate 0.1% w/w Cream | £1.47 |
| Betamethasone Valerate 0.1% w/w Ointment | £1.84 |
| Betamethasone Valerate Ointment (Betnovate RD) | £1.84 |
| Cefixime 400mg (2 x 200mg) | £26.46 |
| Ceftriaxone 1g | £3.62 |
| Ceftriaxone 500 mg (2 x 250mg vials) | £4.60 |
| Chlorphenamine | £2.21 |
| Cilest 63 tablet pack | £4.65 |
| Ciprofloxacin 500mg (2 x 250mg) | £0.31 |
| Clindamycin 300mg bd for 7 days | £17.84 |
| Clindamycin phosphate vaginal cream | £10.86 |
| Clobetasol Propinate (0.05% w/w) Cream (Dermovate) | £2.69 |
| Clobetasol Propionate (0.05% w/w) Ointment | £2.69 |
| Clobetasone Butyrate Cream (Eumovate) | £1.86 |
| Clobetasone Butyrate Ointment (Eumovate) | £1.86 |
| Clobetasone Butyrate, Calcium oxtertracycline & Nystatin Cream (Trimovate) | £12.45 |
| Clotrimazole 100mg Pessary | £0.64 |
| Clotrimazole 200mg Pessary | £1.14 |
| Clotrimazole 500 mg Pessary | £6.99 |
| Clotrimazole Cream 1% | £1.36 |
| Co-Amoxiclav 250/125 (contains PENICILLIN) | £2.03 |
| Co-amxoxiclav 500/125 (contains PENICILLIN) | £2.53 |
| Crotamiton 10% w/w cream | £2.50 |
| Dermol Lotion 500 | £6.04 |
| Desogestrel 75 micrograms | £2.26 |
| Doxycycline 100mg (bd for 14 days) | £3.67 |
| Doxycycline 100mg (bd for 21 days) | £5.51 |
| Doxycycline 100mg (bd for 28 days) | £7.35 |
| Doxycycline 100mg (bd for 7 days) | £2.26 |
| Doxycycline 200mg bd for 4 weeks | £5.51 |
| Emtricitabine 200mg & Tenofovir Disproxil 245mg | £106.00 |
| Emtricitabine 200mg & Tenofovir Disproxil 245mg (3 days) | £10.60 |
| Emulsifying Ointment | £4.82 |
| Erythromycin 250 mg | £8.95 |
| Estradiol 0.5g gel | £5.08 |
| Estradiol 1.0mg gel | £5.85 |
| Estradiol 10 micrograms vaginal tablet | £16.72 |
| Femodene 63 tablet pack | £6.73 |
| Flucloxacillin | £1.41 |
| Fluconazole 150mg | £0.91 |
| Fusidic acid cream | £1.92 |
| GENTAMICIN 240mg for IM injection | £4.13 |
| GYNAEFIX IUD | £27.11 |
| Gardasil 0.5ml - First Dose | £86.50 |
| Gardasil 0.5mls - Second Dose | £86.50 |
| Gardasil 0.5mls - Third Dose | £86.50 |
| Gedarel 20/150 | £5.08 |
| Gedarel 30/150 | £5.08 |
| Hepatitis A & B Combined Vaccine (adult) | £31.18 |
| Hepatitis A Vaccine (2nd at 6 months) | £16.77 |
| Hepatitis A vaccine Day 0 | £16.77 |
| Hepatitis B Vaccine (final at 6 months) | £12.20 |
| Hepatitis B Vaccine - Dose 1 - 10mcg or 20mcg | £12.20 |
| Hepatitis B Vaccine - Dose 2 - 10mcg or 20mcg | £12.20 |
| Hepatitis B Vaccine - Dose 3 - 10mcg or 20mcg | £12.20 |
| Hepatitis B Vaccine - Dose 4 - 10mcg or 20mcg | £12.20 |
| Hepatitis B Vaccine - Extra Dose - 10mcg or 20 mcg | £12.20 |
| Hepatitis B Vaccine 10 mcg 1 month | £12.20 |
| Hepatitis B Vaccine 10 mcg 12 months | £12.20 |
| Hepatitis B Vaccine Day 7 | £12.20 |
| Hepatits B Vaccine Day 21 | £12.20 |
| Hydro-Caine 6mls | £10.50 |
| Hydrocortisone Cream 1% | £1.40 |
| Hydrocortisone Ointment 1% | £1.59 |
| Ibuprofen 200mg | £1.03 |
| Imiquimod 5% | £48.60 |
| Itraconazole 100mg | £3.29 |
| Levonorgestral and Ethinylestradiol 150microgram/30microgram | £2.60 |
| Levonorgestrel 1.5 mg | £3.65 |
| Levonorgestrel 30 micrograms | £0.92 |
| Levosert 52mgs IUS | £66.00 |
| Lidocaine 4% w/w cream | £2.98 |
| Lidocaine 5% m/m Ointment | £8.28 |
| Lidocaine HCL 1% in 2 mls injection | £0.25 |
| Lidocaine HCL 1% in 3.5 mls injection | £0.30 |
| Lidocaine HCL 1% in 5 mls injection | £0.30 |
| Lidocaine HCL 1% in 8mls for IM inj (with IM penicillin) second dose | £0.10 |
| Lidocaine HCL 1% in 8mls for IM injection (with IM penicillin for syphilis) | £0.10 |
| Lidocaine HCL 1% in 8mls for IM injection (with IM penicillin) third dose | £0.10 |
| Lidocaine HCL 2% in 2 mls injection | £0.27 |
| Lidocaine HCL 2% in 5 mls injection | £0.32 |
| Lignocaine 2% Gel | £2.99 |
| Loestrin 20 63 Tablet Pack | £1.99 |
| Loestrin 30 63 Tablet Pack | £1.99 |
| Logynon | £2.60 |
| Marvelon 63 Tablet Pack | £7.10 |
| Mebendazole 100mg | £2.66 |
| Medroxyprogesterone Acetate 104mg in 0.65mls sub cutaneous | £6.90 |
| Medroxyprogesterone Acetate 150mg in 1ml | £6.01 |
| Mefenamic Acid 250mg | £8.17 |
| Mepivacaine Hydrochloride 3% | £0.44 |
| Mepivicaine 3% in 2.2mls | £0.44 |
| Mercilon 63 Tablet Pack | £8.44 |
| Metronidazole 0.75% Vaginal Gel | £4.31 |
| Metronidazole 2g stat dose (400 mg x 5) | £0.52 |
| Metronidazole 400mg (bd for 5 days) | £1.03 |
| Metronidazole 400mg bd for 10 days | £2.07 |
| Miconazole Nitrate 2%w/w, hydrocortisone 1%w/w Cream (Daktocourt) | £2.49 |
| Miconazole Nitrate Cream 20mg/g (Gyno-Daktarin) | £4.33 |
| Miconazole nitrate 20mg per g | £4.33 |
| Millinette 20/75 | £5.41 |
| Millinette 30/75 | £4.12 |
| Mini TT 380 | £12.46 |
| Mirena 52mg IUS | £88.00 |
| Moxifloxacin 400mg od for 10 days | £19.08 |
| Moxifloxacin 400mg od for 14 days | £26.71 |
| Nexplanon 68mg implant | £83.43 |
| Nitrofurantoin 50mg o qds 7 days | £5.08 |
| Nitrofurantoin 50mg o qds for 3 days | £2.18 |
| Nonoxinol-9 | £11.00 |
| Norethisterone 350 micrograms | £2.10 |
| Norethisterone 350 micrograms 84 Tablet Pack | £2.10 |
| Norimin 63 Tablet Pack | £2.28 |
| Nova T 380 | £15.20 |
| Ofloxacin 200mg (one tablet twice daily for 14 days) | £12.54 |
| Ofloxacin 200mg (one tablet twice daily for 7 days) | £6.27 |
| Ofloxacin 200mg (two tablets twice daily for 14 days) | £25.09 |
| Paediatric Hepatitis B Vaccine - Dose 1 - 10mcg | £12.20 |
| Paediatric Hepatitis B Vaccine - Dose 2 - 10 mcg | £12.20 |
| Paediatric Hepatitis B Vaccine - Dose 3 - 10 mcg | £12.20 |
| Paediatric Hepatitis B Vaccine - Dose 4 - 10mcg | £12.20 |
| Paracetamol 500mg | £0.86 |
| Permethrin 5% w/w cream | £8.54 |
| Podophyllotoxin 0.15% Cream | £17.83 |
| Podophyllotoxin 0.5% Solution | £14.49 |
| Raltegravir 400 mg bd for 3 days | £47.14 |
| T- Safe 380A QL | £10.55 |
| TT 380 Slimline | £12.46 |
| Terbinafine Hydrochloride 1% Cream | £2.39 |
| Trimethoprim 200mg | £1.16 |
| Ulipristal Acetate 30mg | £14.05 |
| Xylocaine 1% with adrenaline 1 :200,000 | £1.77 |
