## Supplementary material for "The impact of rapid near-patient STI testing on service delivery outcomes: a controlled interrupted time series study": Table S3

**Table S3**. Intervention-related model estimates for females and males from sensitivity analyses using generalised additive models.

| **Outcome** | **Percent change at time of intervention (95% CI)** | **P-value for post-panther non-linearity of Unity data** |
| --- | --- | --- |
| **MALES – 12^th^ November 2018** |  |  |
| Gonorrhoea culture swabs per consultation | -16.6% (-30.1%, -0.5%) | <0.001 |
| Time to notification | +63.3% (+31.4%, +102.8%) | 0.03 |
| **FEMALES – 29^th^ May 2019** |  |  |
| Gonorrhoea culture swabs per consultation | -11.1% (-29.8%, +12.6%) | <0.001 |
| Time to notification | -14.5% (-34.0%, +10.8%) | <0.001 |
