## Supplementary material for "The impact of rapid near-patient STI testing on service delivery outcomes: a controlled interrupted time series study": Figure S6

**Figure S6.** Modelled estimates of staff capacity for males and females combined. Time trends modelled with splines to allow for non-linearity. All other covariates treated as in the main analysis.

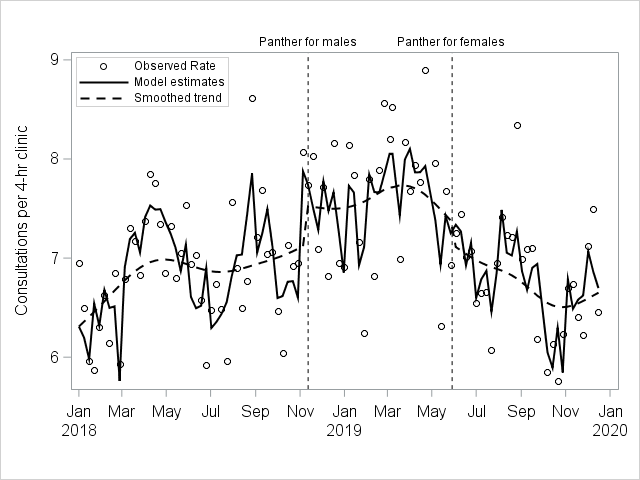
