## Supplementary material for "The impact of rapid near-patient STI testing on service delivery outcomes: a controlled interrupted time series study": Definition S1

**Definition S1.** Criteria used to define complex cases

Criteria for all patients:

a. Patients under 18 years of age
b. Have been/are currently exposed to child sexual exploitation, domestic violence, sexual assault
c. Has a current record of substance misuse
d. Has a current diagnosis of syphilis
e. Has current multiple diagnoses clinical diagnoses (GUMCAD coding B &/or C)
f. Has a history of/current diagnosis of genital herpes or had a swab taken for genital herpes
g. Has had post exposure prophylaxis after sexual exposure to HIV (PEPSE)
h. Needed an interpreter/use of translation service
i. Has current diagnosis of D2B on GUMCAD

Additional criteria for females:

i. Receive contraceptive care
ii. experienced pelvic pain, dyspareunia or post coital bleeding
iii. are pregnant
iv. experienced female genital mutilation.

Additional criteria for males:

v. are bisexual
vi. has sex with men
vii. Experienced testicular pain
viii. has a history/current record of chronic pelvic syndrome
