## Supplementary material for "The impact of rapid near-patient STI testing on service delivery outcomes: a controlled interrupted time series study": Figure S4

**Figure S4.** Modelled outcome estimates for males based on sensitivity analyses using generalised additive models. Both the overall time trend and the post-panther Unity trend were estimated as splines with three degrees of freedom. All other covariates treated as in the main analysis.

| **A.** Gonorrhoea culture swabs (urethral) per consultation | **B.** Median time to notification |
| --- | --- |
| 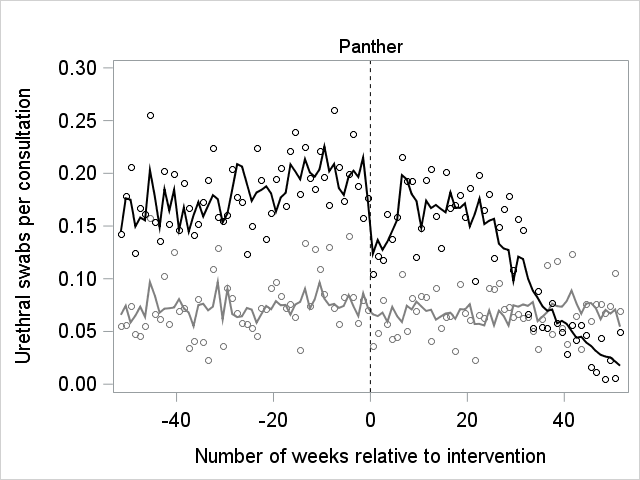 | 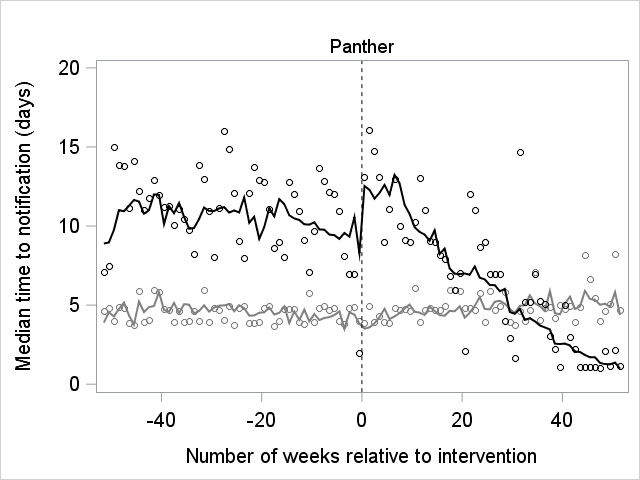 |
| **C.** Examinations per symptomatic attendance | **D.** Follow up attendances per episode |
| 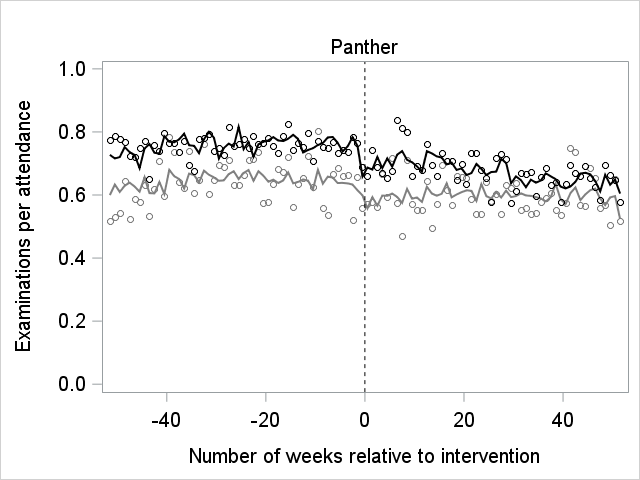 | 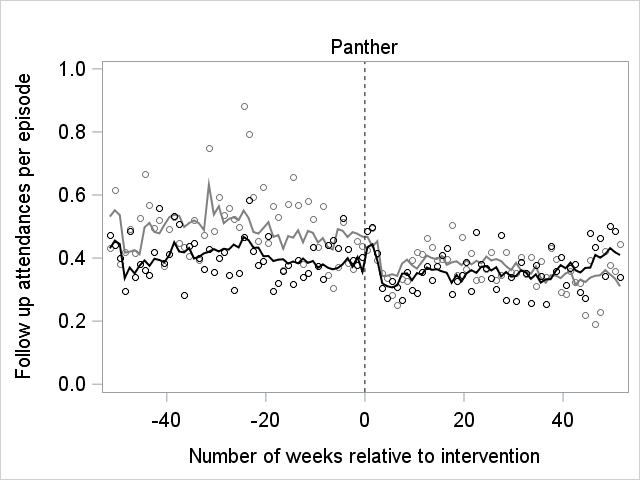 |
| 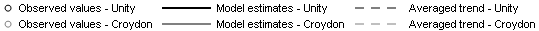 | |
